# ReviewR: A light-weight and extensible tool for manual review of clinical records

**DOI:** 10.1101/2021.10.25.21265151

**Authors:** David A. Mayer, Luke V. Rasmussen, Christopher D. Roark, Michael G. Kahn, Lisa M. Schilling, Laura K. Wiley

## Abstract

**OBJECTIVES:** Manual record review is a crucial step for electronic health record (EHR)-based research, but it has poor workflows and is error prone. We sought to build a tool that provides a unified environment for data review and chart abstraction data entry.

**MATERIALS AND METHODS:** ReviewR is an open-source R Shiny application that can be deployed on a single machine or made available to multiple users. It supports multiple data models and database systems, and integrates with the REDCap API for storing abstraction results.

**RESULTS:** We describe two real-world uses and extensions of ReviewR. Since its release in April 2021 as a package on CRAN it has been downloaded 2,204 times.

**DISCUSSION AND CONCLUSION:** ReviewR provides an easily accessible review interface for clinical data warehouses. Its modular, extensible, and open source nature afford future expansion by other researchers.

**LAY SUMMARY:** When doing research using data from electronic health records (EHRs), data may need to be extracted by hand, either to perform the study or to ensure its accuracy. However many researchers can’t access the EHR for this purpose. Even when researchers have access, they must flip between their review list, the EHR, and the location they are recording the results of their review, which is difficult and can cause errors. We developed a software application, ReviewR, to make this process easier and less error prone and have used it in two real-world projects.

## BACKGROUND AND SIGNIFICANCE

Electronic health records (EHRs) are commonly used for clinical, translational, and population health research.^1–3^ Although significant advances have been made in methods for automated data extraction, chart/record review (i.e., extracting or abstracting information from a patient’s EHR manually) remains a crucial tool for EHR-based research.^4–6^ However manual chart review is complex, time consuming, and error prone.^7^ Typically chart review occurs in the source EHR (e.g., Epic, Cerner) with the extraction results stored in a secondary system (e.g., Excel, REDCap^8^). This leaves researchers juggling multiple application windows which has poor usability and increases the potential for data entry errors (like mistyping the patient ID or entering data on the wrong row of the spreadsheet). Further, many EHRs lack sophisticated search capabilities (e.g., using regular expressions to search for multiple relevant terms simultaneously), increasing the complexity of record review. Providing researchers with a single, integrated, and optimized tool would help improve the efficiency and quality of manual record review.

Although traditionally chart review has relied upon using the source EHR, some institutions restrict EHR access to only clinically credentialed staff. Others may require researchers to access deidentified data warehouses or restrict access to only those patients covered under research agreements. This prevents researchers from performing traditional chart review because researchers are restricted to using a data warehouse or specific data extracts. Thus there is a need for a tool to support record review that uses data from clinical data warehouses instead of the traditional EHR interface. A number of tools have been developed to support visualization and searching of clinical data warehouses including i2b2,^9^ OHDSI’s Atlas,^10^ Leaf,^11^ and EMERSE (for clinical notes only),^12^ however these tools do not support integrated chart abstraction. Additionally these tools all require server-based deployment which typically requires support from data warehouse and/or IT teams. Tools exist for text annotation including the Clinical Language Annotation, Modeling, and Processing Toolkit (CLAMP),^13^ Knowtator,^14^ and the brat rapid annotation tool,^15^ however these tools are designed to work with individual notes or sentences rather than entire patient records. Given that manual record review often relies on a combination of structured and unstructured data from the EHR, there is an unmet need for a lightweight and flexible tool that has a unified review interface.

## OBJECTIVES

Our primary objective was to design and build a tool that provides a streamlined workflow for performing manual chart review from clinical data warehouses. Specifically, the tool should have 1) search and filter capabilities that can be applied across an entire patient record, and 2) a unified interface that minimizes context switching by supporting simultaneous review of data and data entry into a chart abstraction form. In the development of the tool, we also followed three key design principles: 1) adhere to electronic data capture best practices, 2) support lightweight and portable deployment options that allow use by individual researchers or organizations, 3) provide easy extensibility to support different clinical data models and relational database management systems (RDBMS) used to store and query clinical data.

## MATERIALS AND METHODS

ReviewR is an R Shiny^16^ application built using modules and the {golem} framework^17^ for production grade app development.^18^ This approach allows ReviewR to be distributed as a regular R package supporting R v3.5.0 or later.^19^ ReviewR was developed and documented using {devtools},^20^ the {tidyverse},^21^ and a number of other utilities.^22–27^ The user interface leverages multiple dashboarding and widget toolkits^28–31^ and CSS/JavaScript tools.^32,33^ Clinical records are presented using the {DT} package,^34^ an R interface to the DataTables JavaScript library,^35^ which allows users to easily filter columns and perform complex searches using regular expressions. Support for multiple RDBMSs is provided through {dbplyr}^36^ - a package that converts regular dplyr-based code into SQL using a {DBI}^37^ mediated database connection. ReviewR currently supports clinical data stored in SQLite,^38^ PostgreSQL,^39^ and Google BigQuery,^40,41^ but can be extended to include any RDBMS supported by {dbplyr}.

Data models are supported in ReviewR using a schema definition (i.e., list of table and associated column names) and a matched set of display functions. The data model is automatically detected by selecting the schema with the highest number of table/column name matches. We provide development functions for extending ReviewR to support any custom data model and offer out of the box support for multiple versions of the OMOP common data model^42^ and the MIMIC3 database.^43^ Chart abstractions are captured in REDCap^8^ with ReviewR translating REDCap instruments into native Shiny widgets and transmitting data using the REDCap API and associated R interfaces.^44,45^ ReviewR supports multiple instruments and a number of commonly used field types and data validation rules (see **Table 1**).

**Table 1.** Supported REDCap Field and Data Validation Types.

| Field Type | Validation Options |
| --- | --- |
| Checkboxes (Multiple Answers) | NA |
| Multiple Choice - Drop-down List (Single Answer) | NA |
| Multiple Choice - Radio Buttons (Single Answer) | NA |
| Notes Box (Paragraph Text) | NA |
| Text Box (Short Text, Number, Date/Time, ...) | Date (M-D-Y); Integer |
| True - False | NA |
| Yes - No | NA |

To test the extensibility and face validity of ReviewR, we conducted two demonstration projects. First, we sought to extend ReviewR to connect to a new RDBMS (Microsoft SQL Server) to support phenotype-based chart review. Second, we sought to extend ReviewR to support a custom data model and compare chart review conducted using ReviewR to chart review performed in the source EHR. One of the authors (CDR) reviewed 50 records in both Epic and ReviewR using a crossover design (i.e., 25 records were reviewed first in Epic and then ReviewR, with the other 25 reviewed in Reviewer first) to extract history of intracranial aneurysm, subarachnoid hemorrhage (SAH), and aneurysmal SAH. We then analyzed review concordance.

## RESULTS

ReviewR can be used locally by individual users, or it can support multiple concurrent users when deployed to a Shiny Server. Docker images for both local and server installations may be built from the Dockerfiles included in the package source. ReviewR is available for download on CRAN (https://cran.r-project.org/package=ReviewR) and is distributed with the open source 3-Clause BSD License on GitHub (https://github.com/thewileylab/ReviewR/). Users can also trial ReviewR without download using ShinyApps.io (https://thewileylab.shinyapps.io/ReviewR/). All versions include a SQLite demonstration database that includes ten records from the CMS SynPUF dataset and “clinical notes” from a medical transcription training corpus.^46–48^

### Using ReviewR

Users access the Setup tab to connect ReviewR to their patient database (**Figure 1**). Users select the RDBMS from the drop down menu which opens the connection module. Completing this connection sets the user into *view mode* where they can explore patient records. Optionally, users can enter *review mode* that supports data abstraction by connecting to and configuring REDCap. Users must have created a REDCap project and instrument through their institution and have access to an API key for the project. Once connected, users are prompted to configure the instrument which allows ReviewR to automatically populate both the patient and reviewer names in the instrument, reducing errors and repetitive data entry tasks. After connecting, users select records to review from the “Patient Search” tab. This tab displays a list of patients, demographics, and abstraction status (if configured). Users can click on the patient identifier to navigate to the individual’s record.

**Figure 1.**
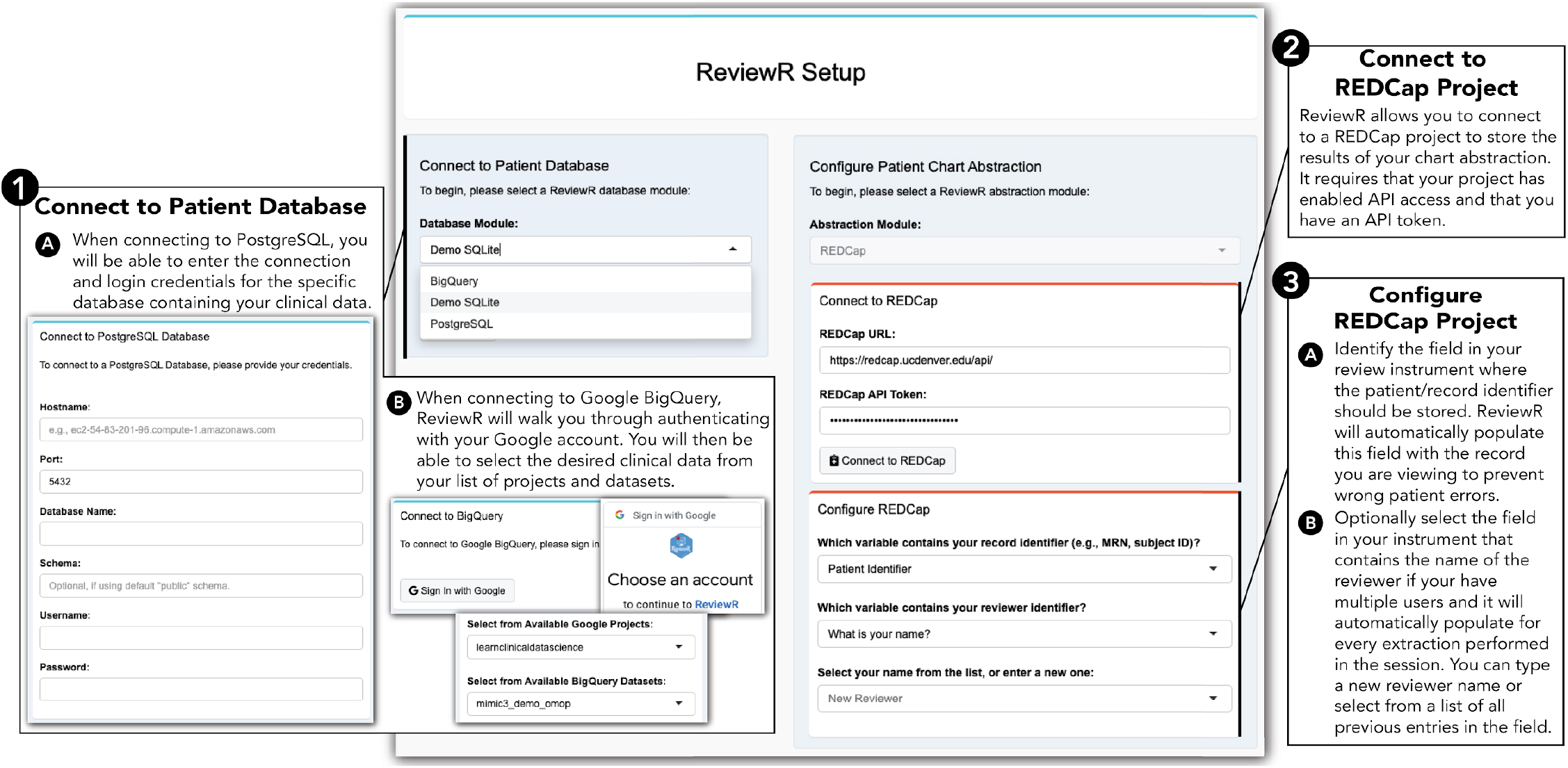
ReviewR Setup Tab. This page allows users to enter the view mode by connecting to their patient database first selecting the appropriate RDBMS and then providing connection credentials (for Postgres) or using the “Sign in to Google” interface (for Bigquery). Optionally users can enter review mode by connecting to a previously created REDCap project using an API key and then configuring the connection to identify the review field that will hold the patient and (optionally) reviewer identifiers.

Individual patient records are displayed in the “Chart Review” tab (**Figure 2**). Patient demographic information is always available for the selected patient at the top of the screen, along with navigation options to progress through the patients. Below the demographic data is a tabbed interface containing the information from each table in the connected database. Users can easily search this data using regular expressions both within and across columns, tabs/tables, and even records, filtering to only the relevant entries with matches highlighted for easy identification. When abstraction is configured, the REDCap abstraction instrument will be displayed to the right of the patient data. Data is uploaded to REDCap using the “Save to REDCap” button, with ReviewR warning users if it detects that previously entered data has been changed.

**Figure 2.**
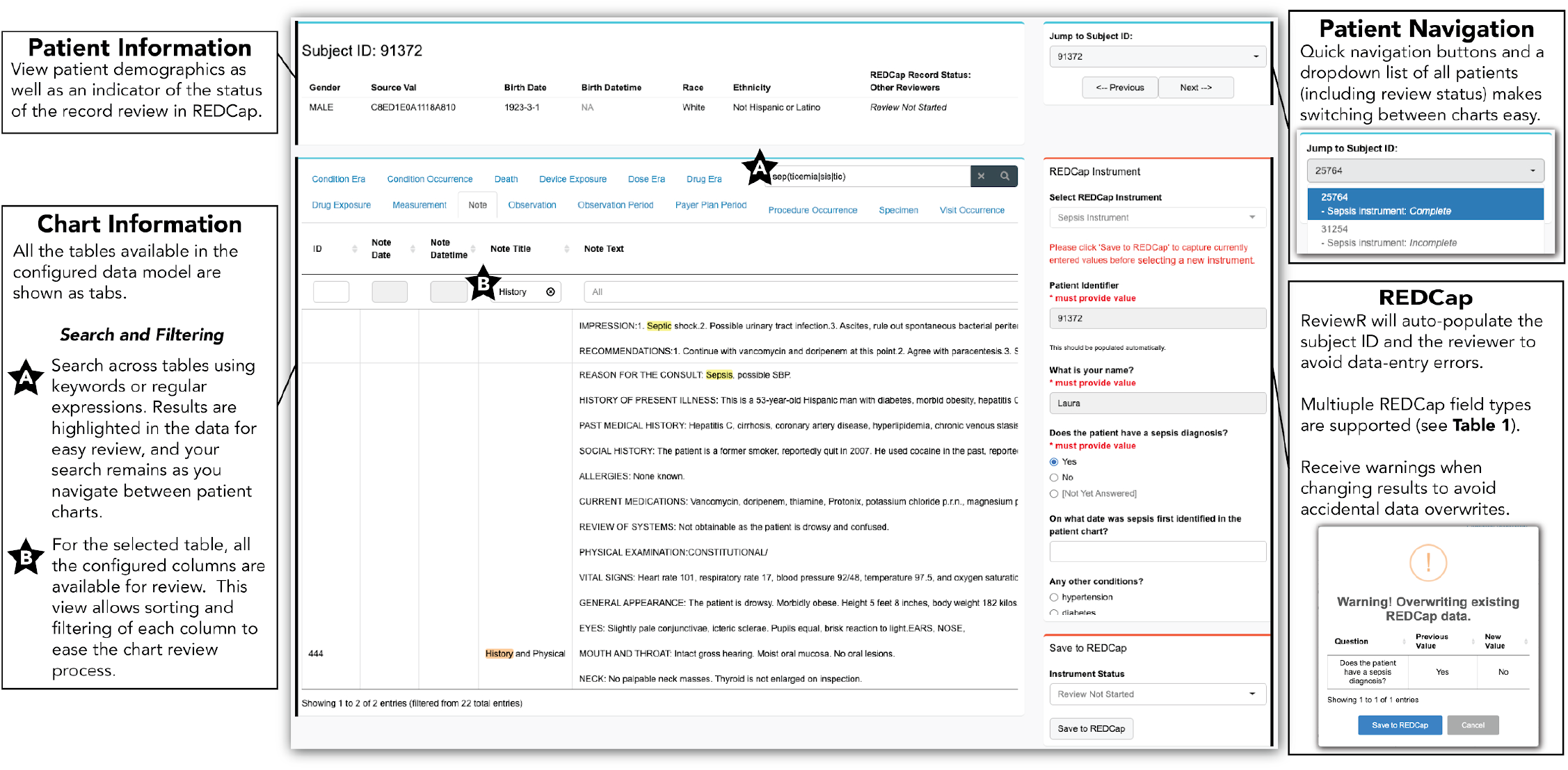
ReviewR Chart Review Tab. This page allows users to review the medical chart for a single patient. The patient information is shown at the top, including the chart review status as recorded in REDCap. Chart information shows all of the available tables for the configured data model. Users can perform a search (including the use of regular expressions) across all of the tables, and results will be highlighted (A). For each of the columns in a data table, users can do additional filtering and sorting (B). Users are guided through charts using quick navigation controls, and the configured REDCap instrument is displayed alongside the chart data.

### Extending ReviewR

We have included a number of developer functions to help users extend ReviewR to support additional data models and RDBMSs. Adding a new data model requires limited R programming experience and is outlined in **Figure 3**. Users provide a data model schema as a CSV file and then select the table and fields containing patient information. More technical users may optionally customize table displays by editing the ReviewR generated R code. Adding a new RDBMS requires more programming expertise as users must develop a Shiny module that captures the database credentials and returns a valid {DBI} connection. However, ReviewR offers functions and templates to support this process. Vignettes describing this process and full package documentation are available at https://reviewr.thewileylab.org.

**Figure 3.**
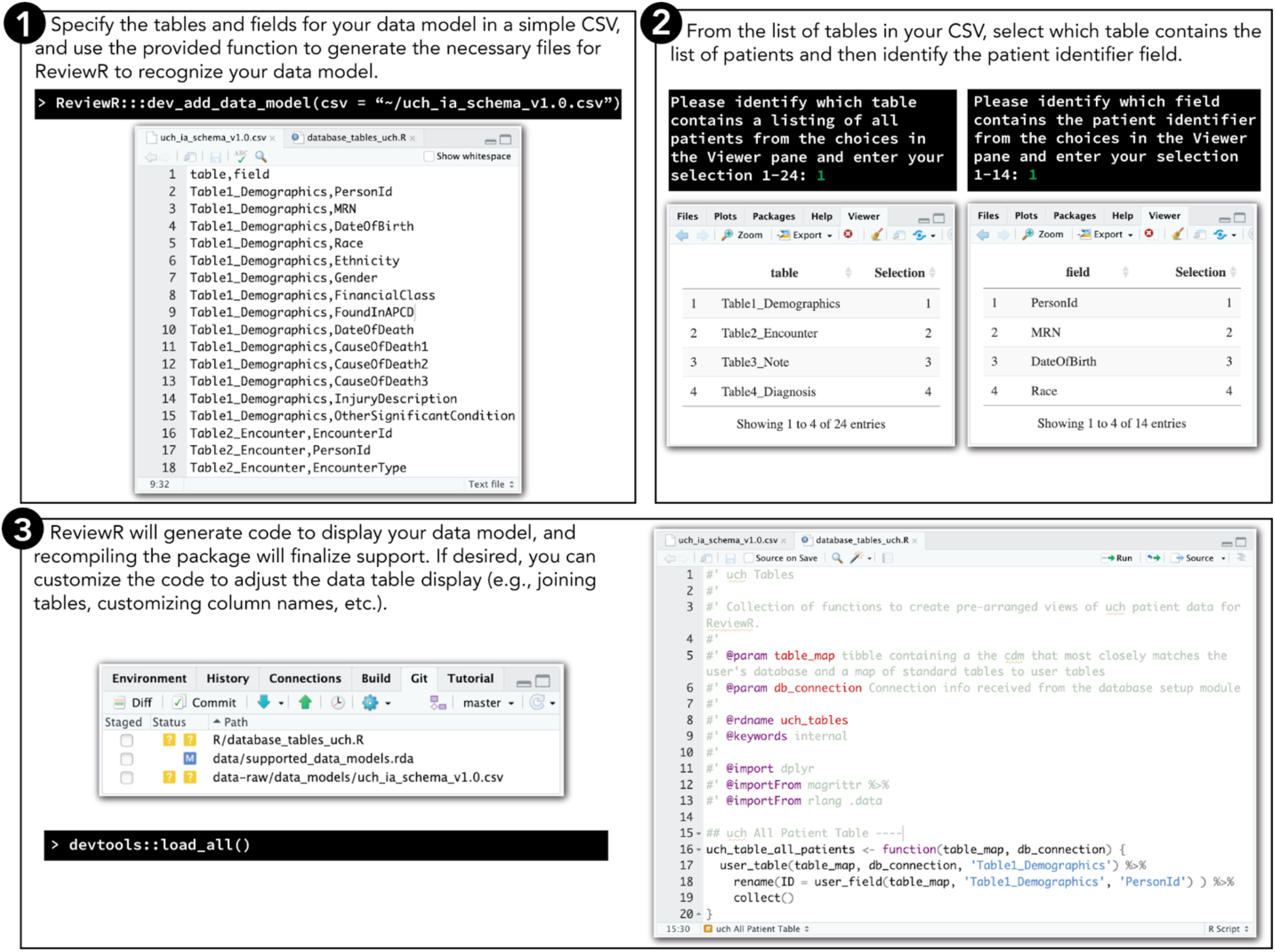
ReviewR Process for Adding Support for a Custom Data Model. Users can easily add support for a new data model using build in developer tools. Step 1: The user must create a schema file that contains all tables and associated field names as a csv file. Within the R console they pass this file to the ‘dev_add_data_model()’ function. Step 2: ReviewR will walk users through selecting the table containing patient demographics (used to define the “Patient Search” tab) and the column name containing the patient identifier (used to auto populate the REDCap instrument). Step 3: ReviewR adds the schema to the list of supported data models and generates template R code for displaying the database as is. Reloading the package finalizes this support. More technical users may customize the R code to change table displays, join tables (for example the OMOP data model requires joining to the concept table to have an informative information display), etc.

### Demonstration Projects

The first demonstration project was performed at Northwestern University which restricts EHR access for clinical use only. Researchers needed to perform chart review to confirm the case/control status of a random subset of patients identified by a heart failure phenotyping algorithm. LVR successfully built a {DBI} connection object to connect ReviewR to the Northwestern Medicine Enterprise Data Warehouse OMOP instance which is stored in Microsoft SQL Server. The results of this chart review were subsequently published.^49^

The second demonstration project was performed at the University of Colorado to test the face validity of chart review conducted in ReviewR compared to the source EHR and to build a gold-standard cohort for aneurysm phenotype algorithm development. Although 50 records were identified for review, one record was accidentally skipped during ReviewR extraction and was excluded from our analysis. There was an overall concordance rate of 94% (**Table 2**), with ReviewR identifying two patients with a history of an aneurysm and one patient with a history of SAH that were not identified during chart review in Epic.

**Table 2.** Second Demonstration Project Review Results.

| Extraction Order | Concordant | Discordant | Total |
| --- | --- | --- | --- |
| ReviewR -> Epic | 23 | 2<sup>†</sup> | 25 |
| Epic -> ReviewR | 23 | 1<sup>‡</sup> | 24 |
| **Total** | 46 | 3 | 49 |

<sup>†</sup>ReviewR identified one additional aneurysm and aneurysmal SAH compared to Epic review
<sup>‡</sup>ReviewR identified one additional aneurysm compared to Epic review

In addition to these known uses, ReviewR has been downloaded from CRAN 2,315 times between April 2, 2021 and October 16, 2021.

## DISCUSSION

To our knowledge, ReviewR represents the first open source tool explicitly designed to support manual record review that provides a unified environment for data review and chart abstraction data entry. ReviewR was designed to meet multiple research challenges that we have observed within our research experience. First and foremost, the unified interface provides researchers with a streamlined workflow while automated entry of patient and reviewer identifiers target common sources of data entry errors and inefficiencies. Second, ReviewR fills gaps present at many institutions where research access to source EHRs is restricted or researchers only have access to de-identified data warehouses. In many cases these institutions also lack record-level display tools making manual record review challenging and laborious. ReviewR’s lightweight design and ability for standalone deployment by individual researchers helps bridge these gaps.

A key strength of ReviewR comes from it’s extensible design and use of Shiny modules. By partitioning the application into a modular structure, adding support for a new data model or RDBMS requires little to no modification of ReviewR core code. Further the inclusion of developer functions to streamline, and extensive documentation to support, this process will hopefully attract a community of users who can help grow the functionality of ReviewR. Additionally, these modules can be easily repurposed by other researchers in their own applications. For example the database modules all provide generic connection objects that are broadly useful.

Finally our demonstration projects prove that ReviewR can be deployed in practice to support chart review across institutions and use cases. Moreover, our paired testing of ReviewR compared to Epic-based chart review helps support the face validity of performing chart review using clinical data warehouses. Although these warehouses typically contain only a portion of the patient record, in our testing ReviewR actually had a higher sensitivity, identifying three more cases than the source EHR. It is possible this is due to the advanced search capabilities of ReviewR that surfaced notes that otherwise might have been missed within the source EHR.

However, ReviewR does have limitations. First, the user interface for ReviewR has not been formally evaluated or studied. Although anecdotally our experience has been positive, a more formal assessment is needed before any claims could be made about gains in efficiency. Second, ReviewR has a limited number of databases and data models that it currently supports, and only connects to data warehouses and not EHR systems (though with the development of R packages to support FHIR-based exchange this may become possible). Finally, although R is widely adopted, the selection of R Shiny as the application platform for ReviewR may preclude its deployment as an enterprise-wide solution where R Shiny is not currently supported. By allowing ReviewR to run in multiple modes (standalone, hosted, Docker container), we hope that potential users will find a secure deployment option that works for their organization.

## CONCLUSION

In this paper, we describe the development and implementation of ReviewR - a flexible and extensible tool that can be used to perform chart abstractions from EHR data stored in clinical data warehouses. Its unified user interface streamlines the researcher workflow and can reduce potential errors during the review process. As a free and open source solution, we hope its continued adoption and refinement can improve the speed and quality of manual record review.

## Data Availability

All code for ReviewR is publicly available on both GitHub and CRAN.

https://CRAN.R-project.org/package=ReviewR

https://github.com/thewileylab/ReviewR/

https://reviewr.thewileylab.org/

## CONTRIBUTORS

LKW conceived the idea of ReviewR, acquired funding, and supervised the overall direction and scope of the project. LKW and LVR developed an initial prototype of the software; DM developed the release version of ReviewR and drafted all software documentation. LVR and CR performed beta testing, and MGK and LMS provided substantive input on design and functionality of ReviewR. LKW and DM developed the initial draft of this manuscript, and all authors reviewed and approved the final manuscript.

## ACKNOWLEDGEMENTS

We would like to thank the Google Cloud Healthcare team; attendees at the 2018 OHDSI Collaborator Showcase, 2019 AMIA Informatics Summit, and 2021 R/Medicine conference; the University of Colorado Informatics Interest Group; the Colorado Center for Personalized Medicine; and the Northwestern Biomedical Informatics and Data Science Group for helpful comments on interim presentations of this work.

